# The genetic architecture of postoperative delirium after major surgery and its relationship with non-postoperative neurocognitive conditions

**DOI:** 10.1101/2025.09.01.25334224

**Authors:** Richard A. Armstrong, Paul Yousefi, Ben Gibbison, Golam M Khandaker, Tom R Gaunt

**Author notes:** **Correspondence to:** Richard A Armstrong **Email:** **X:** @drrichstrong.

## Abstract

**Background:** Postoperative delirium is the most common postoperative complication in older individuals. Genome-wide association studies (GWAS) can provide insights into how genetic factors influence postoperative risk. We examined the genetic architecture of postoperative delirium after major surgery and its relationship with related cognitive conditions (delirium of any type and Alzheimer’s disease, including the *APOE* ε4 allele).

**Methods and Findings:** A case-control GWAS was performed in UK Biobank to identify genetic variants associated with postoperative delirium, adjusted for age, sex, genetic chip and the first ten principal components. These results were then used in genetic correlation and polygenic risk score analyses to investigate shared genetic risk between postoperative delirium and a) delirium of all causes, and b) Alzheimer’s disease.

The GWAS (1016 cases, 139,148 controls) identified seven Single Nucleotide Polymorphisms (SNPs) that mapped to four genes (*APOE, TOMM40, APOC1* and *PVRL2*); p <5 x 10^-8^. Five SNPs remained significant after excluding pre-existing dementia, and two after excluding subsequent dementia. The lead SNP was rs429358, a missense variant of *APOE*. Genetic correlation and polygenic risk score analyses revealed evidence of shared genetic architecture and risk between postoperative delirium and Alzheimer’s disease (rho 0.68, 95% CI [0.46,0.81]; p<0.001). After adjustment for age and sex, the *APOE* ε4 isoform had a dose-response effect on risk (odds ratios for one and two copies: 1.75, 95% CI [1.53,2.0], and 4.19, 95% CI [3.25,5.41], respectively; p <0.001). The main limitations of the study include the reliance upon clinical coding for outcome definition and limited statistical power to detect small or modest genetic effects.

**Conclusions:** We identified genetic variants associated with increased risk of postoperative delirium. We also found evidence of shared genetic liability with Alzheimer’s disease via *APOE*, complementing recent large-scale studies in all-cause delirium. If validated the findings have potential clinical applications including preoperative risk stratification and early identification of pre-clinical Alzheimer’s disease risk.

**Author Summary:** *Why was this study done?:*

- Postoperative delirium is the most common postoperative complication in older individuals.
- Little is known about the genetic basis of postoperative delirium.
- Knowledge of the genetic risk factors could help treatment or prevention in the future.

*What did the researchers do and find?:*

- We performed a genome-wide association study of postoperative delirium including 1016 cases and 139,148 controls.
- We found genetic variants associated with increased postoperative delirium risk.
- We also found genetic similarity between postoperative delirium, all-cause delirium, and Alzheimer’s disease.

*What do these findings mean?:*

- Postoperative delirium has shared biology with Alzheimer’s disease and may represent unmasking of pre-clinical disease.
- Individuals with genetic risk for Alzheimer’s disease have increased risk of postoperative delirium and vice versa.
- Due to the relatively small number of cases and the fact that we may not have detected all the true positive diagnoses in the dataset, we were unable to detect small or modest genetic effects in this study.

## Introduction

Postoperative delirium is the most common postoperative complication in older individuals, with an incidence of up to 65% [1,2]. It has substantial health and resource implications including increased morbidity, mortality and healthcare costs [1,3]. Hypotheses of the underlying mechanisms span a range of pathways including release of excitotoxic neurotransmitters, neuronal injury and inflammation, physiological stressors, metabolic derangements, electrolyte disorders and genetic factors [1,2,4–8]. Postoperative delirium has been associated with an increased risk of cognitive decline and incident dementia [9] but there is conflicting evidence on how the acute occurrence of postoperative delirium interacts with longer-term cognitive decline and neurodegeneration [10].

Whilst the aging global population and increasing numbers of older adults undergoing surgery have made perioperative brain health an increasing focus of research and clinical initiatives [11], the ability of clinical teams to produce personalised assessments of clinical risk for individuals remains limited. Improved detection of underlying biological risk in conjunction with established clinical risk factors could aid in preoperative risk prediction, facilitate better-informed shared-decision making and allow targeting of multicomponent interventions to prevent delirium [11] with potential reductions in morbidity, mortality and resource utilisation.

Population-based genome-wide association studies (GWAS) are a powerful approach for identifying genetic risk factors associated with complex conditions of multifactorial aetiology. Their results can aid prediction but also identify underlying biological mechanisms and potential targets for intervention. They have been extensively applied to a wide range of clinical phenotypes [12], but rarely to postoperative delirium specifically. Previous GWAS that have included postoperative neurocognitive complications have failed to identify any genome-wide significant associations [13–15] - most likely because they suffered from limited sample sizes and heterogeneity of outcomes.

This study aimed to evaluate the association between genetic variants genome-wide and postoperative delirium following major surgery, and to explore whether postoperative delirium shares genetic risk factors with related conditions, specificially delirium of all causes and Alzheimer’s disease. The limitations of previous studies are addressed by defining a major surgery cohort in a large scale national biobank and focusing on a single outcome phenotype.

## Methods

### Study populations

The data source for the primary analysis was UK Biobank, a long-term prospective cohort study that provides a large-scale biomedical database and research resource containing genetic, lifestyle and health information from half a million UK participants recruited between 2006 and 2010 (http://www.ukbiobank.ac.uk). The UK Biobank study was approved by the North-West Multi-centre Research Ethics Committee and all participants provided written informed consent [16]. Additionally, publicly available GWAS summary statistics from the FinnGen study were used as an external cohort for genetic correlation analyses. FinnGen is a large-scale genomics initiative that has analysed over 500,000 Finnish biobank samples and correlated genetic variation with health data to understand disease mechanisms and predispositions. The project is a collaboration between research organisations and biobanks within Finland and international industry partners [17].

### Genotyping

UK Biobank genotyping was performed by Affymetrix using two purpose-designed arrays. Genotype data were quality controlled, phased and ∼96 million genotypes were imputed using the Haplotype Reference Consortium and UK10K haplotype resources. FinnGen genotyping used a ThermoFisher Axiom custom chip array with imputation to a reference dataset derived from Finnish whole-genome sequences. Further information on the methods and QC pipelines are available [16,18].

### Inclusion and exclusion criteria

Eligible participants were those undergoing major, inpatient surgery after their date of enrolment in UK Biobank. Linked hospital inpatient data is available for all UK Biobank participants with follow-up to 31 May 2022 for Wales, 31 August 2022 for Scotland and 31 October 2022 for England [19]. Major surgery was defined by Office of Population Censuses and Surveys Classification of Interventions and Procedures, Version 4 (OPCS-4) code using the Bupa Schedule of Procedures and Abbott classification [20,21] and diagnoses of delirium were identified using International Classification of Diseases, Tenth Revision (ICD-10) code F05. Exclusion criteria included planned day case surgery and a previous diagnosis of delirium (see S2 Appendix for more detail on phenotype definition and exclusions).

### Outcome definition

The primary outcome was a first diagnosis of delirium within 30 days of surgery, including diagnoses made during readmission. Controls were defined as those undergoing major surgery with no delirium diagnosis within 30 days. The 30-day window was used to align with recommendations on the nomenclature of postoperative delirium [22] and other widely-used sources [23].

### Genome-wide Association Study (GWAS)

Case–control GWAS adjusted for age, sex, genetic chip and the first ten principal components was performed using regenie (v3.2.2) [24]. Participants with missing data for these variables were excluded. Quality control and ancestry definition were performed by previously established workflows and methods [25]. Post-hoc analyses were conducted using Functional Mapping and Annotation of Genome-Wide

Association Studies (FUMA GWAS [26]). Independent significant Single Nucleotide Polymorphisms (SNPs) were defined as those meeting a genome-wide significance threshold of *p* < 5 x 10^-8^ and independent of each other at r^2^ < 0.6. Lead SNPs were identified from independent significant SNPs at r^2^ < 0.1. Gene-based analysis was performed using Multi-marker Analysis of GenoMic Annotation (MAGMA [27]).

### Sensitivity analyses

The primary GWAS analysis did not include additional clinical covariates as they may be mediators of genetic risk or colliders given the selection of a surgical patient cohort. To explore the robustness of the primary analysis the following sensitivity analyses were performed: a) after excluding participants who had a previous dementia diagnosis; b) excluding participants who had a previous dementia diagnosis or were diagnosed with dementia after the index procedure; c) stratified by cardiothoracic/non-cardiothoracic surgery; d) adjusted for Charlson Comorbidity Index (0-1 vs ≥2, [28]); and e) conditional on Apolipoprotein E (*APOE*) ε4 haplotype count (0, 1 or 2) to identify associated variants independent of *APOE*.

### Post-GWAS follow-up analyses

Data-driven exploratory follow-up analyses were informed by the results of the primary GWAS analysis. Two approaches were taken to formally assess shared genetic architecture between postoperative delirium and a) delirium of any type; b) Alzheimer’s Disease: genetic correlation analysis and polygenic risk scoring.

### Genetic correlation analysis

Genetic correlation analyses were performed between the UKBiobank GWAS results presented here and the external FinnGen study cohort. GWAS summary statistics (Freeze 9, https://r9.finngen.fi/) were obtained for a) delirium (any cause); and b) Alzheimer’s disease. Local Analysis of [co]Variant Association (LAVA [29]) was used to analyse bivariate local genetic correlation between these phenotypes and postoperative delirium (using the summary statistics from the primary GWAS analysis in UK Biobank). The genome is partitioned into 2,495 blocks for LAVA analysis so a Bonferroni-adjusted p-value of 0.05/2,495 (2 x 10^-5^) was used to define statistical significance.

### Polygenic risk score analysis

Polygenic risk scores (PRS) calculate a weighted sum of risk based on alleles associated with a disease or outcome phenotype. The Polygenic Score Catalog (https://www.pgscatalog.org/) was used to identify polygenic risk scores for Alzheimer’s disease. No polygenic risk scores were identified for delirium. The *pgsc_calc* workflow [30] was used to download, variant match and calculate individual polygenic scores for the primary UK Biobank cohort. Individual scores were then centred and scaled before being summed and averaged. Odds ratios for the association between increasing quintile of polygenic risk score for Alzheimer’s disease and postoperative delirium were estimated by logistic regression with adjustment for age and sex using the *glm* function in R (version 4.4.0). Sensitivity analyses were performed with additional adjustment for Charlson Comorbidity Score (0-1 vs ≥2, [28]). A Bonferroni correction for the number of comparisons against the reference quintile was applied, resulting in a significance threshold of p < 0.0125 (0.05/4).

### Apolipoprotein E (APOE) genotype analysis

It has been recommended that the *APOE* region of chromosome 19 should be excluded from Alzheimer’s disease polygenic risk scores, with the *APOE* ε4 allele included as an additional covariate in modelling, due to its outsize effect relative to other regions across the genome [31]. The subset of polygenic scores identified above which did not include the *APOE* region (defined as a 10 kilobase window either side of region from the start position of *TOMM40*, chr19:45,394,477, to the stop position of *APOC1*, chr19:45,422,606, build 37/hg19 [31]) were therefore considered in an additional analysis, adjusted for age, sex and *APOE* ε4 allele count (0, 1, 2).

This study is reported as per the Strengthening the Reporting of Observational Studies in Epidemiology (STROBE) guideline (S1 Checklist).

## Results

### Participant characteristics

A total of 1016 cases of postoperative delirium and 139,148 controls were identified (Figure 1). Individuals who developed postoperative delirium tended to be older, male and had higher prevalence of comorbidities (Table 1). Compared to controls, a higher proportion of patients in the postoperative delirium group underwent emergency (532 (52%) vs 29,530 patients (21%)) and cardiothoracic surgery (172 (17%) vs 5041 patients (3.6%)).

**Figure 1.**
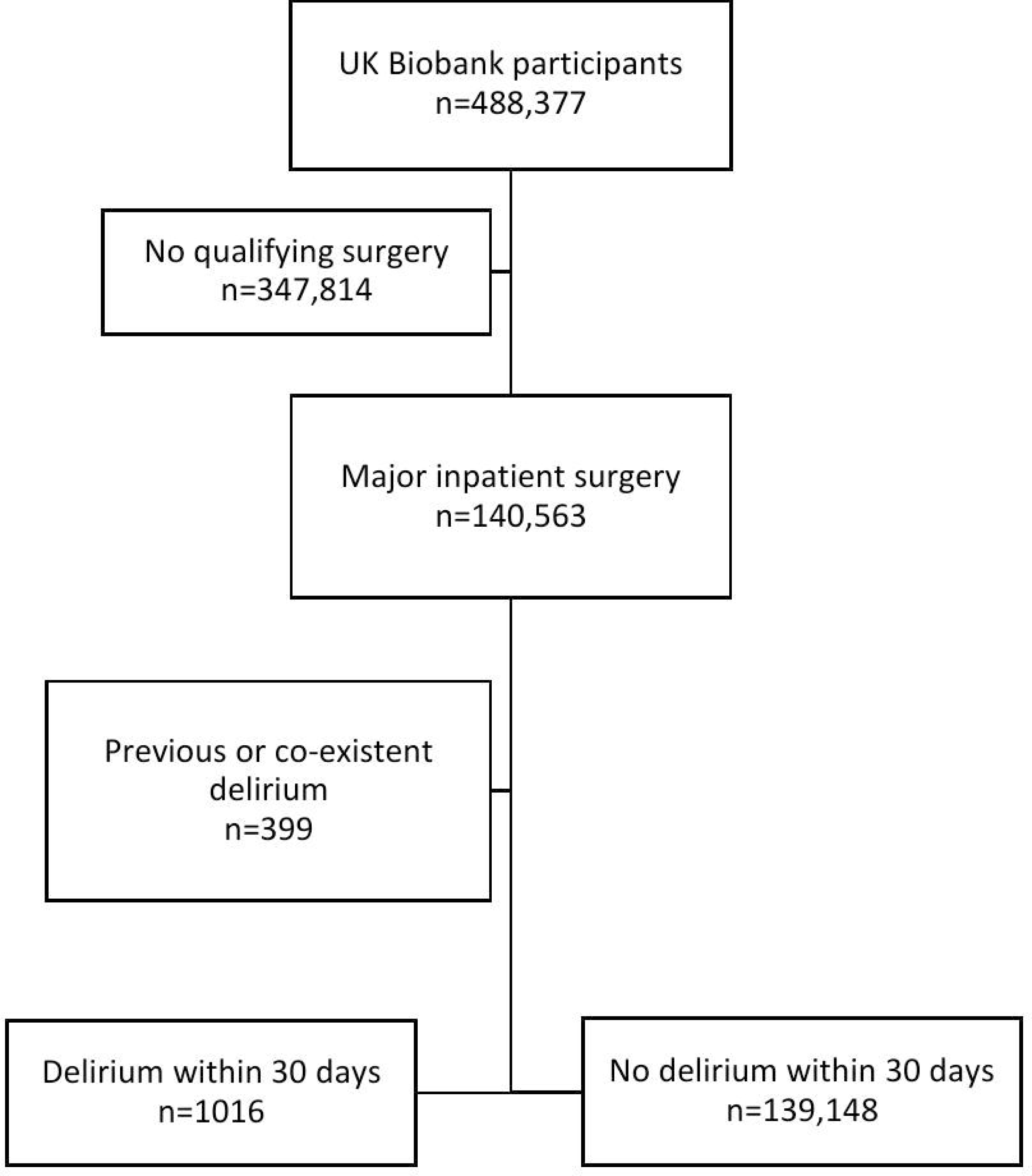
Study flow diagram.

**Table 1.** Participant characteristics for postoperative delirium cases and controls.

|  | **Cases**<br>**n = 1016** | **Controls**<br>**n = 139,148** | **p-value** |
| --- | --- | --- | --- |
| Age (yr), mean (SD) | 72 (6) | 64 (8) | <0.001 |
| Sex, number (%) |  |  | <0.001 |
| Female | 384 (38) | 77,244 (56) |  |
| Male | 632 (62) | 61,904 (44) |  |
| Ethnicity, number (%) |  |  | 0.2 |
| Asian or Asian British | 9 (0.9) | 2301 (1.7) |  |
| Black or Black British | 10 (1.0) | 1644 (1.2) |  |
| Chinese | 0 (0) | 239 (0.2) |  |
| Mixed | 5 (0.5) | 675 (0.5) |  |
| Other ethnic group | 3 (0.3) | 984 (0.7) |  |
| White | 978 (97) | 132,676 (96) |  |
| Missing | 11 (1.1) | 629 (0.5) |  |
| Charlson Comorbidity Index, median (IQR) | 1 (0–3) | 0 (0–1) | <0.001 |
| Missing | 25 (2.5) | 15,817 (11) |  |
| Admission method, number (%) |  |  | <0.001 |
| Elective | 434 (43) | 106,725 (77) |  |
| Emergency | 532 (52) | 29,530 (21) |  |
| Maternity | 0 (0) | 113 (<0.1) |  |
| Not known | 0 (0) | 29 (<0.1) |  |
| Transfer | 50 (4.9) | 2751 (2.0) |  |
| Surgical specialty, number (%) |  |  | <0.001 |
| Cardiothoracic surgery | 172 (17) | 5041 (3.6) |  |
| General surgery | 196 (19) | 26,436 (19) |  |
| Neurosurgery | 55 (5.4) | 5192 (3.7) |  |
| Other | 186 (18) | 36,532 (26) |  |
| Trauma and orthopaedics | 338 (33) | 53,722 (39) |  |
| Urology | 37 (3.6) | 10,628 (7.6) |  |
| Vascular | 32 (3.1) | 1597 (1.1) |  |

Numbers are number (proportion), mean (standard deviation, SD) or median (interquartile range, IQR). Continuous variables compared using Wilcoxon rank sum test; categorical variables using Pearson's Chi-squared test or Fisher's exact test for cell counts of 5 or fewer. Additional details can be found in Table A in S3 Appendix.

### Genome-wide significant associations for postoperative delirium

Seven independent, statistically significant SNPs were identified within a single genetic risk region on chromosome 19 (Figure 2, Table 2). The Q-Q plot showed no evidence of systematic inflation (genomic inflation factor (λ) 1.01, Figure 3). The lead SNP was rs429358, a missense variant in the *APOE* gene that, together with rs7412, defines the ε2/ε3/ε4 isoforms linked to Alzheimer’s disease risk. A total of 21 genes were mapped to this region, including four (*APOE*, *APOC1, TOMM40* and *PVRL2*) that were significantly associated in MAGMA gene-level analysis (Figure 4; Table B, S3 Appendix). The Q-Q plot showed no evidence of systematic inflation (genomic inflation factor (λ) 1.0, Figure 5).

**Figure 2.**
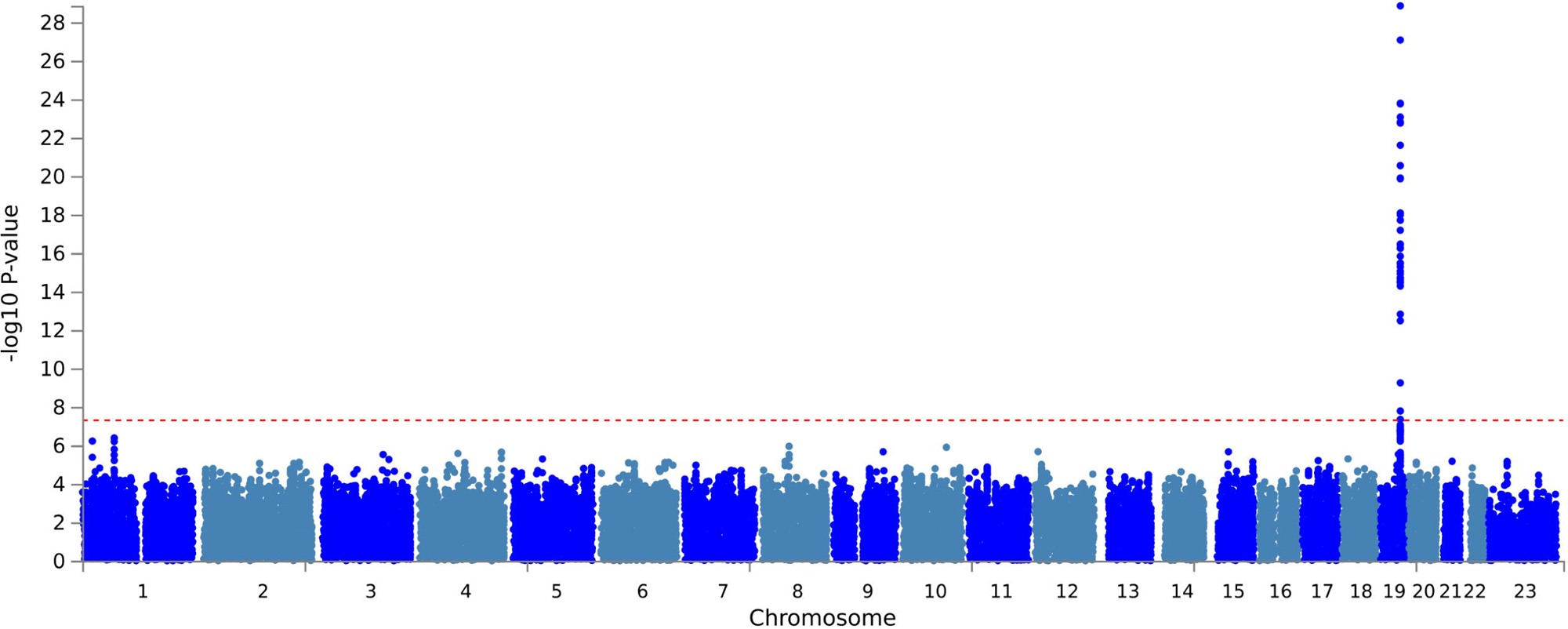
Manhattan plot of case-control GWAS of postoperative delirium results (adjusted for age, sex, chip and first ten principal components). The red line represents a genome-wide significant p value of 5 x 10^-8^.

**Figure 3.**
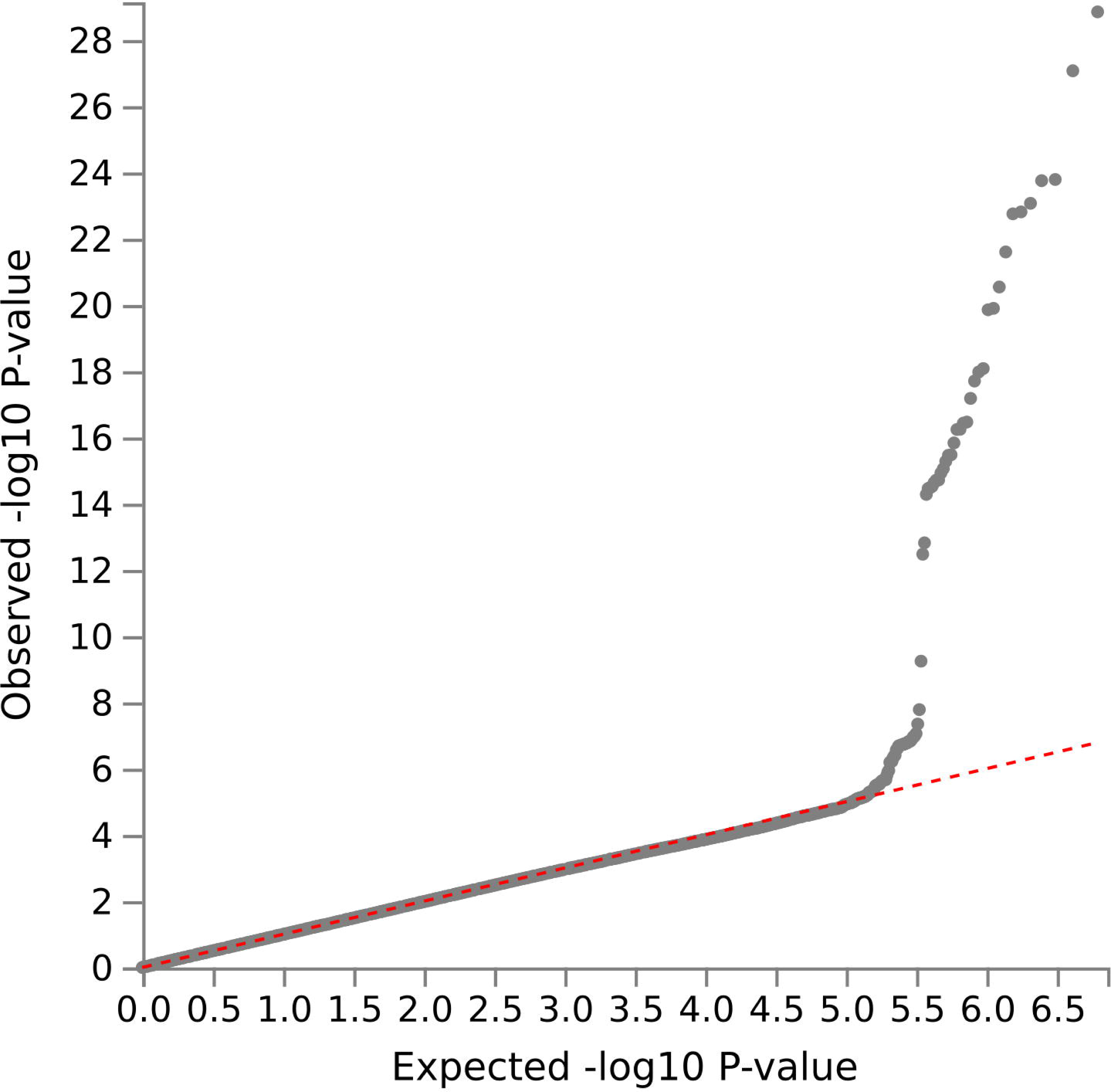
Q-Q plot of case-control GWAS of postoperative delirium results (primary analysis). Genomic inflation factor (λ) 1.01.

**Figure 4.**
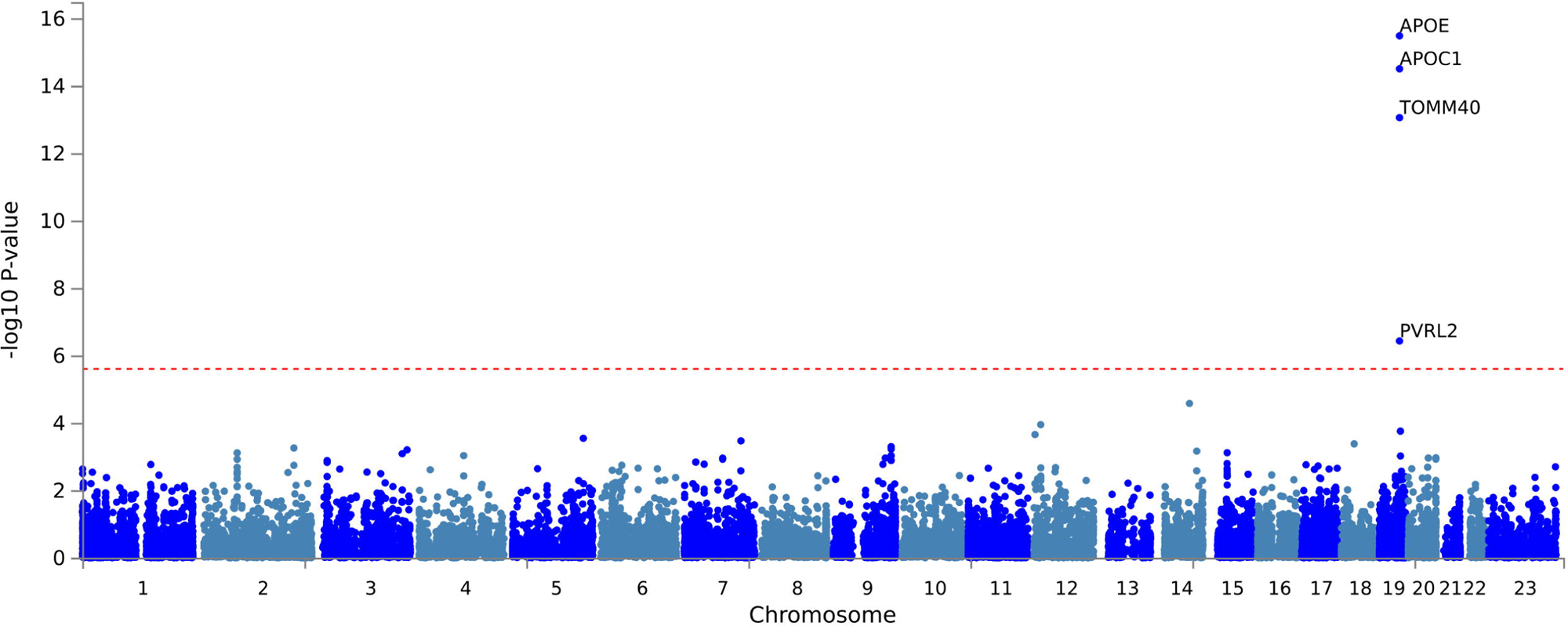
Manhattan plot for MAGMA gene testing from case-control GWAS of postoperative delirium (adjusted for age, sex, chip and first ten principal components). The red line represents a genome-wide significant p value of 2.5 x 10^-6^.

**Figure 5.**
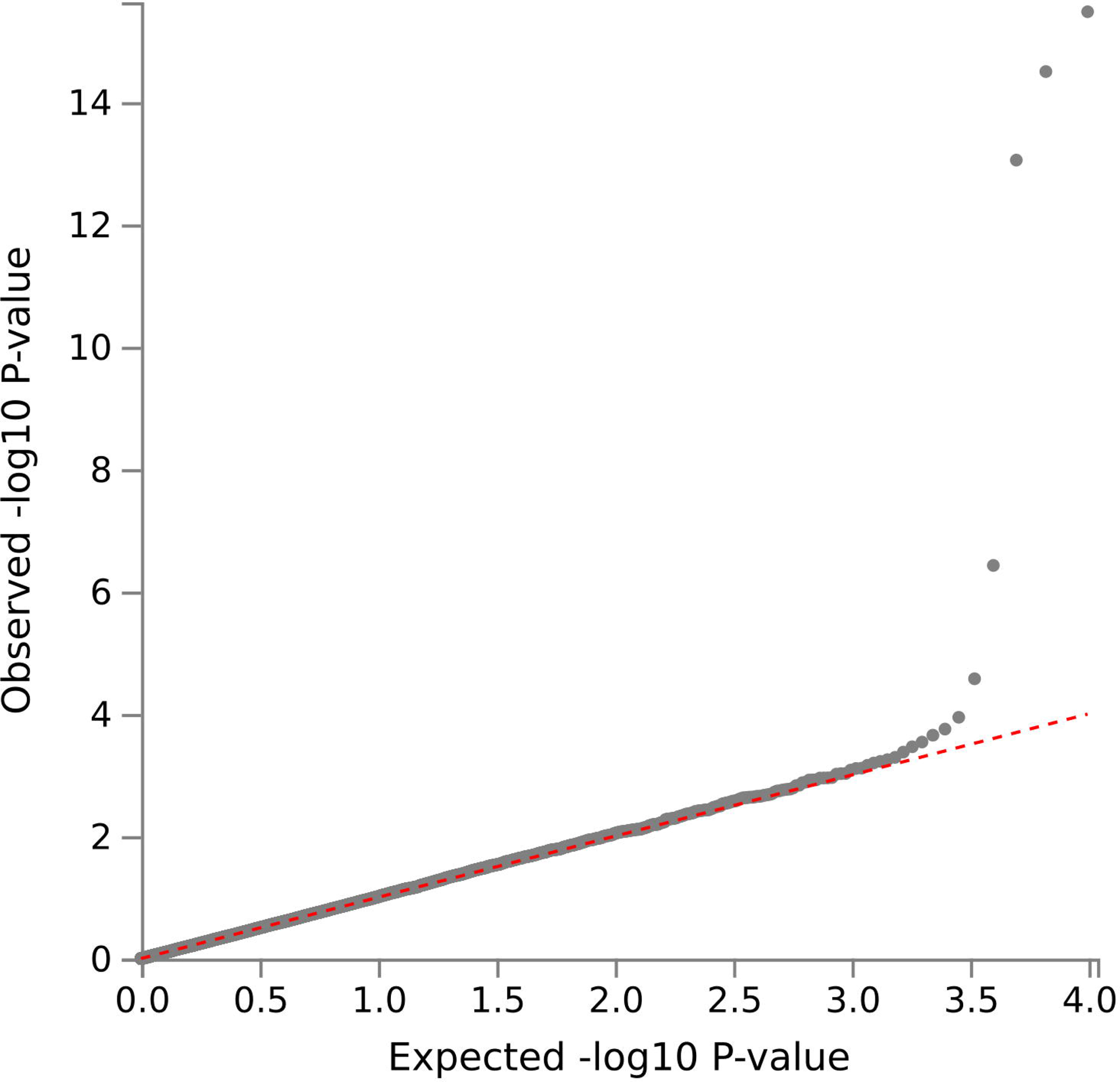
Q-Q plot for MAGMA gene testing from case-control GWAS of postoperative delirium results (primary analysis). Genomic inflation factor (λ) 1.0.

**Table 2.** Independent significant single-nucleotide polymorphisms passing genome-wide significance threshold in case-control GWAS of postoperative delirium.

| rsID | Chr:position | Nearest gene | Function | Effect allele | Odds ratio (95% CI) | p |
| --- | --- | --- | --- | --- | --- | --- |
| rs429358 | 19:45411941 | APOE | exonic | T | 0.54 (0.49,0.59) | $1.41 \times 10^{-29}$ |
| rs157592 | 19:45424514 | APOC1 | intergenic | A | 0.58 (0.53,0.64) | $1.77 \times 10^{-23}$ |
| rs157582 | 19:45396219 | TOMM40 | intronic | C | 0.65 (0.59,0.71) | $3.42 \times 10^{-17}$ |
| rs11556505 | 19:45396144 | TOMM40 | exonic | C | 0.62 (0.56,0.69) | $3.33 \times 10^{-16}$ |
| rs10119 | 19:45406673 | TOMM40 | UTR3 | G | 0.7 (0.64,0.77) | $3.34 \times 10^{-13}$ |
| rs75627662 | 19:45413576 | APOE | downstream | C | 0.72 (0.65,0.79) | $5.66 \times 10^{-10}$ |
| rs12691088 | 19:45418486 | APOC1 | Intronic | G | 0.45 (0.35,0.58) | $1.65 \times 10^{-8}$ |

rsID: Reference SNP cluster ID; UTR3: 3' untranslated region.

### GWAS results after excluding existing or incident dementia

After excluding individuals with a pre-existing diagnosis of dementia (56 cases (5.7%) and 375 controls (0.3%)), 5 of the 6 independent significant SNPs remained significant. However, rs75627662 (a downstream variant of *APOE*) no longer reached genome-wide significance. Three genes, *APOE*, *APOC1* and *TOMM40*, remained significant on MAGMA testing (Figures A and B, S3 Appendix). When patients who went on to be diagnosed with dementia during UK Biobank follow-up were also excluded (141 cases, 13.9%, and 2358 controls, 1.7%), two SNPs remained genome-wide significant, rs429358 and rs157592. The MAGMA results were unchanged (Figures C and D, S3 Appendix).

### GWAS results in cardiothoracic and non-cardiothoracic surgery groups

The results for the non-cardiothoracic surgery subgroup (798 cases, 126,973 controls) were similar to those in the overall cohort with five independent significant SNPs identified in the same risk locus on chromosome 19 and three of the four genes remained significant on MAGMA testing (*APOE*, *APOC1* and *TOMM40*). The lead SNP was the same as in the primary GWAS and the other variants either overlapped with the primary analysis or were in strong linkage disequilibrium (r^2^ > 0.97) with them (Table C in S3 Appendix). The cardiothoracic analysis was likely underpowered and returned no genome-wide significant results at *p* < 5 x10^-8^. Multiple SNPs reached a lower indicative threshold of significance including the lead SNP from the primary analysis, rs429358 (p = 3.8 x 10^-7^; Table D in S3 Appendix).

### GWAS results adjusted for Charlson Comorbidity Index

A total of 495 (48.7%) cases and 26,659 (19.2%) controls had a Charlson Comorbidity Index (CCI) ≥2 (χ^2^ = 562.42, p < 0.001). GWAS results after adjustment for CCI were consistent with the primary analysis, returning the same seven independent significant SNPs on chromosome 19 (Table E in S3 Appendix).

### GWAS results conditional on Apolipoprotein E (APOE) ε4 allele count

*APOE* ε*4* allele counts differed between cases and controls (0, 1 and 2 copies in 566 (57.4%), 348 (35.3%) and 71 (7.2%) cases vs 95,489 (71.6%), 34,754 (26.1%) and 3,082 (2.3%) controls; χ^2^ = 159.37, p < 0.001). After adjustment for *APOE* ε4 allele count, all the independent significant SNPs from the primary GWAS analysis lost statistical significance (Figure E in S3 Appendix).

### Genetic correlation with Alzheimer’s disease

Significant local genetic correlation was identified on chromosome 19 between postoperative delirium and both delirium of all causes and Alzheimer’s disease (Table 3). The correlated region (chr19:45040933-45893307) encompassed the independent significant SNPs from the primary GWAS.

**Table 3.** Results of Local Analysis of [co]Variant Association (LAVA) testing between postoperative delirium GWAS results and FinnGen GWAS summary statistics.

| FinnGen phenotype | FinnGen sample size (cases / controls) | Rho (95% CI) | R<sup>2</sup> (95% CI) | p |
| --- | --- | --- | --- | --- |
| Delirium (all) | 3,039 / 356,660 | 0.63<br>(0.46,0.81) | 0.40<br>(0.22,0.66) | 3.7 x 10<sup>-10</sup> |
| Alzheimer's disease | 13,393 / 363,884 | 0.68<br>(0.57,0.80) | 0.47<br>(0.32,0.65) | 5.2 x 10<sup>-21</sup> |

Rho represents the local genetic correlation coefficient. R<sup>2</sup> represents the proportion of variance in postoperative delirium explained by the comparator phenotype.

### Association of genetic predisposition for Alzheimer’s disease with postoperative delirium

Prior to averaging, forty-two PRS for Alzheimer’s disease were identified and passed variant matching (Table F in S3 Appendix). Increased polygenic risk for Alzheimer’s disease was associated with increased risk of postoperative delirium in the UK Biobank cohort after adjusting for age and sex (adjusted OR PRS quintile 4: 1.49 (95% CI [1.2,1.85]), PRS quintile 5: 2.29 (95% CI [1.87,2.8]); p <0.001; Table 4).

**Table 4.** Odds ratio for postoperative delirium by Alzheimer’s disease polygenic risk score (PRS) quintile. Results are shown for all 42 polygenic risk scores, adjusted for age and sex, and the subset which did not include the *APOE* region (15 *APOE*-independent scores), adjusted for age, sex and *APOE* ε4 genotype.

| Term | All scores (n=42) |  |  | *APOE*-independent scores (n=15) |  |  |
| --- | --- | --- | --- | --- | --- | --- |
|  | Unadjusted odds ratio (95% CI) | Adjusted odds ratio (95% CI) | P value | Unadjusted odds ratio (95% CI) | Adjusted odds ratio (95% CI) | P value |
| PRS quintile 1 | 1.00 (reference) | 1.00 (reference) | - | 1.00 (reference) | 1.00 (reference) | - |
| PRS quintile 2 | 1.06 (0.85,1.34) | 1.07 (0.85,1.35) | 0.54 | 1.23 (1,1.52) | 1.24 (1,1.53) | 0.052 |
| PRS quintile 3 | 1.25 (1,1.56) | 1.25 (1,1.57) | 0.047 | 1.32 (1.07,1.62) | 1.32 (1.07,1.62) | 0.01 |
| PRS quintile 4 | 1.46 (1.18,1.81) | 1.49 (1.2,1.85) | <0.001 | 1.24 (1,1.53) | 1.24 (1,1.53) | 0.049 |
| PRS quintile 5 | 2.18 (1.79,2.66) | 2.29 (1.87,2.8) | <0.001 | 1.46 (1.19,1.79) | 1.50 (1.22,1.84) | <0.001 |
| Age | - | 1.22 (1.2,1.23) | <0.001 |  | 1.22 (1.20,1.23) | <0.001 |
| Sex - male | - | 1.82 (1.6,2.07) | <0.001 |  | 1.82 (1.60,2.07) | <0.001 |
| *APOE* ε4 x1 | - | - | - | | 1.75 (1.53,2.00) | <0.001 |
| *APOE* ε4 x2 | - | - | - | | 4.19 (3.25,5.41) | <0.001 |

These results were largely unchanged in a sensitivity analysis adjusting for Charlson Comorbidity Index (Table G in S3 Appendix).

### Effect of APOE genotype on the association of Alzheimer’s disease risk with postoperative delirium

Fifteen of the polygenic risk scores identified for Alzheimer’s disease contained no variants in the *APOE* region (*APOE*-independent scores, Table F in S3 Appendix). The effect size of increasing PRS quintile was reduced compared to *APOE*-inclusive scores and the presence of one or more *APOE* ε4 isoforms was associated with increased risk of postoperative delirium (Table 4). Similar results were seen with adjustment for Charlson Comorbidity Index (Table G in S3 Appendix).

## Discussion

In this study, using a large national biobank with linked healthcare data, we identified a genetic risk locus for postoperative delirium on chromosome 19 which included seven independent significant SNPs and four mapped genes meeting genome-wide significance. Follow-up analyses in an independent cohort confirmed the presence of shared genetic risk between postoperative delirium and Alzheimer’s disease.

Previous GWAS which have included postoperative delirium as an outcome have failed to find any genome-wide significant results, but were likely underpowered [13–15]. Our study, to the best of our knowledge the largest GWAS of postoperative delirium to date, is substantially better powered than previous analyses and implicates *APOE* – an established genetic determinant of Alzheimer’s disease – in postoperative delirium risk. The lead SNP identified (rs429358) remained significant when patients with a pre-existing or subsequent diagnosis of dementia were excluded, as did the significance of the *APOE* gene on MAGMA testing. We have identified additional genes associated with postoperative delirium (*APOC1*, *TOMM40* and *PVRL2*), which were previously reported to be associated with a range of neurocognitive phenotypes including dementia, cognitive function and decline, brain biomarkers and cerebral imaging metrics (Table B and Figure F, S3 Appendix).

However, a sensitivity GWAS conditional on *APOE* ε4 allele count resulted in the loss of significance of all SNPs in the *APOE* region suggesting *APOE* is the sole contributing factor at this genetic risk locus. We did not replicate the results of an earlier candidate gene study that identified an association between the cholinergic genes *CHRM2* and *CHRM4* and postoperative delirium [14] (all three identified variants showed null effects, Table H in S3 Appendix). However, we used a genome-wide approach which has methodological advantages including increased genetic coverage, reduced bias and lower false positive rates and therefore represents a more robust analysis [32].

Post-GWAS follow-up analyses showed that genetic predisposition for Alzheimer’s disease is associated with risk of perioperative delirium, including a dose-response effect of *APOE* ε4 allele count. Previous studies investigating the association between *APOE* ε4 and postoperative delirium have shown mixed results. A recent study of 19,331 patients (2086 of whom had postoperative delirium) found *APOE* ε4 carriers to be at increased risk of neurocognitive disorders compared with non-carriers [33]. However, the neurocognitive traits in this study were defined using Phecodes – which include a wider range of medical diagnoses than the more specific ones used in our study [34] – and the authors did not report the timepoint at which outcomes were assessed. Several other studies have found no association [35–37] whilst one small study (29 cases of postoperative delirium) did report increased risk with the ε4 allele [38].

Taken together, these data suggest shared genetic architecture between postoperative delirium, delirium in general, and Alzheimer’s disease via *APOE*. This is supported by a large-scale genetic meta-analysis of all-cause delirium which identified *APOE* as a strong delirium risk factor and showed that incorporating ε4 status into prediction models improved incident delirium prediction [39]. The study identified a further four risk loci for delirium which we did not replicate in our postoperative delirium cohort, likely due to differences in statistical power and analytical approach. We did, however, find that *APOE*-independent PRS for Alzheimer’s disease were associated with postoperative delirium, mirroring the smaller contribution made by other genomic regions. A role for APOE-related pathways in postoperative neurocognitive outcomes has biological plausibility through effects on neuroinflammation and blood-brain barrier integrity [40]. In the perioperative setting, *APOE* ε4 carriers have shown greater postoperative decreases in functional connectivity in key Alzheimer’s disease risk regions [41]. Further validation of these findings may lead to potential clinical applications including preoperative risk stratification based on associated variants or existing polygenic risk scores (e.g. for Alzheimer’s disease) and the identification of potential therapeutic targets for the reduction of postoperative delirium risk. Alternatively, the acute occurrence of postoperative delirium might represent an unmasking of underlying neurocognitive vulnerability or an undiagnosed chronic condition, e.g. pre-clinical Alzheimer’s disease. If genetic liability for Alzheimer’s disease extends to postoperative delirium, patients found to be at increased risk of delirium may benefit from additional monitoring long-term.

The strengths of this study include the use of a large prospective cohort with linked healthcare data that is subject to rigorous quality control and validation procedures. We defined our exposure using published criteria and employed robust statistical methods to deal with the large case-control imbalance. We were also able to extend our findings through a range of post-GWAS analyses using an independent validation cohort. The study does have some limitations. The diagnosis of delirium relied upon ICD-10 clinical coding and was not prospectively screened for. Validation studies have shown that ICD-10 coding in administrative databases has low sensitivity but excellent specificity for postoperative delirium compared to prospective clinical screening [42,43]. As such there will be some misclassification bias, particularly true cases which are included in the control group. This will likely have attenuated the observed genetic effects and reduced our statistical power to detect small or modest effects. Whilst is also possible that some cases of delirium preceded surgery, the risk of this was minimised as far as possible. It is also possible that some cases in the 30-day follow-up period were related to other factors, for example infection or hospitalisation, but these occurred in the context of recovery from major surgery so were included in line with recommendations on the nomenclature of postoperative cognitive changes. Whilst UK Biobank represents a large cohort of patients there is a recognised “healthy volunteer” selection bias [44] and so the results may not be entirely generalisable to the wider UK population. Any secondary data analysis relying on routinely-collected administrative data is limited by the fields available, with some potentially relevant factors such as the American Society of Anesthesiologists (ASA) physical status grade, surgical urgency and type of anaesthesia not recorded. Finally, whilst rates of some comorbidities and surgical characteristics differed between cases and controls, these variables may lie on the causal pathway or act as selection-related colliders in a major surgical cohort. We therefore did not adjust for them in the primary GWAS but examined their influence through a range of sensitivity analyses which support the robustness of our findings.

In conclusion, we have used a large national biobank with linked healthcare data to identify seven SNPs and four genes associated with increased risk of postoperative delirium. Data from an independent cohort suggests there is shared genetic architecture and risk between postoperative delirium, delirium more broadly, and

Alzheimer’s disease. If validated, these findings have potential clinical benefit through risk stratification based on genotype or polygenic risk score data and early identification of future Alzheimer’s disease risk in the perioperative setting.

## Authors’ contributions

RAA: study design, data analysis, preparing first and subsequent drafts of the paper; PY: study design, drafting paper; BG: study design, drafting paper; GK: study design, drafting paper; TG: study design, drafting paper.

## Supporting information

S1 STROBE checklist

## Data Availability

• The individual-level participant data for the primary GWAS and PRS analyses are available from UK Biobank via their controlled access process (https://www.ukbiobank.ac.uk/use-our-data/). FinnGen GWAS summary statistics are available from the FinnGen website (https://www.finngen.fi/en).
Author-generated code to support the analyses is available on Github (https://github.com/raarmstrong/gwas-postop-delirium/; DOI:10.5281/zenodo.18196799). GWAS summary statistics for the primary analysis will be available from the data.bris Research Data Repository (DOI TBC).

## Acknowledgements

This research has been conducted using the UK Biobank Resource under Application Number 128619. This includes linked data provided by patients and collected by the NHS as part of their care and support and data assets made available by National Safe Haven as part of the Data and Connectivity National Core Study, led by Health Data Research UK in partnership with the Office for National Statistics and funded by UK Research and Innovation (grant ref MC_PC_20058).

Quality Control filtering of the UK Biobank data used the process described by R.Mitchell, G.Hemani, T.Dudding, L.Corbin, S.Harrison, L.Paternoster[25].

This work was carried out using the computational and data storage facilities of the Advanced Computing Research Centre, University of Bristol (http://www.bristol.ac.uk/acrc/).

## Competing interests

I have read the journal’s policy and the authors of this manuscript have the following competing interests: TRG receives funding from GlaxoSmithKline, Biogen and Roche for unrelated research. The other authors have declared that no competing interests exist.

## Financial disclosure

This study was funded by a Wellcome Trust GW4-CAT PhD Programme for Health Professionals PhD Fellowship awarded to RAA [316275/Z/24/Z]. RAA, PY, GMK and TRG are supported by the Medical Research Council Integrative Epidemiology Unit at the University of Bristol (RA, TG: MC_UU_00032/3; PY: MC_UU_00032/4; GMK: MC_UU_0032/6). GMK acknowledges additional funding from the Wellcome Trust (grant numbers: 201486/Z/16/Z and 201486/B/16/Z), the Medical Research Council (grant numbers: MR/W014416/1; MR/S037675/1; MR/Z50354X/1; and MR/Z503745/1. PY, TRG, and GMK are also supported by the UK National Institute for Health and Care Research (NIHR) Bristol Biomedical Research Centre (grant number: NIHR 203315). The views expressed are those of the authors and not necessarily those of the UK NIHR or the Department of Health and Social Care. The funders had no role in study design, data collection and analysis, decision to publish, or preparation of the manuscript.

## Supporting Information files and legends

**S1 Checklist. Strengthening the Reporting of Observational Studies in Epidemiology (STROBE) checklist**

**s2 Appendix. Supplementary methods**

**S3 Appendix. Tables and figures**

**Table A** Additional participant characteristics for case-control GWAS of postoperative delirium. Comorbidities assessed at time of surgery. Operative categories are listed as Abbott/Bupa classification.

**Table B** Mapped genes from primary case-control GWAS of postoperative delirium. minGwasP represents the minimum p-value associated with the gene; IndSigSNPs lists the independent significant SNPs tagged to that gene.

**Table C** Independent significant SNPs in non-cardiothoracic surgery case-control GWAS of postoperative delirium (sensitivity analysis). rsID: Reference SNP cluster ID.

**Table D** Top 10 SNPs from case-control GWAS of postoperative delirium in cardiothoracic cohort (sensitivity analysis). rsID: Reference SNP cluster ID.

**Table E** Independent significant single-nucleotide polymorphisms passing genome-wide significance in case-control GWAS of postoperative delirium after adjustment for age, sex, chip, first 10 PCs and Charlson Comorbidity Index (sensitivity analysis). rsID: Reference SNP cluster ID.

**Table F** Polygenic risk scores from PGS Catalog passing matching and included in polygenic risk score analyses. *APOE-independent scores.

**Table G** Odds ratio for postoperative delirium by Alzheimer’s disease polygenic risk score (PRS) quintile. Results are shown for all 42 polygenic risk scores, adjusted for age and sex, and the subset which did not include the APOE region (15 APOE-independent scores), adjusted for age, sex, APOE ε4 genotype and Charlson Comorbidity Index (0-1 vs >=2).

**Table H** Results from primary analysis (case-control GWAS of postoperative delirium) for variants identified in candidate gene study by Heinrich et al. rsID: Reference SNP cluster ID.

**Figure A** Manhattan plot for case-control GWAS of postoperative delirium after previous dementia diagnoses excluded. The red line represents a genome-wide significant p value of 5 x 10^-8^.

**Figure B** Manhattan plot for MAGMA testing for case-control GWAS of postoperative delirium after previous dementia diagnoses excluded.

**Figure C** Manhattan plot for case-control GWAS of postoperative delirium after previous or subsequent dementia diagnoses excluded. The red line represents a genome-wide significant p value of 5 x 10^-8^.

**Figure D** Manhattan plot for MAGMA testing for case-control GWAS of postoperative delirium after previous or subsequent dementia diagnoses excluded.

**FIGURE E** Manhattan plot for case-control GWAS of postoperative delirium conditional on APOE ε*4* allele count (adjusted for age, sex, chip, first 10 PCs).

**Figure F** GWAS catalog reported genes for case-control GWAS of postoperative delirium (primary analysis).

## Notes

### Competing Interest Statement

TRG receives funding from GlaxoSmithKline, Biogen and Roche for unrelated research. The other authors declare that they have no conflict of interest.

### Author Declarations

UK Biobank (http://www.ukbiobank.ac.uk) and Finngen (https://r9.finngen.fi).

### Summary of Updates

Author accepted version. Abstract updated to include main covariates and number formatting; main limitations added to abstract and author summary; formatting of CIs changed; 2 supplementary figures moved into main text.

