## Supplementary material for "The genetic architecture of postoperative delirium after major surgery and its relationship with non-postoperative neurocognitive conditions": S2 Appendix

**Supplementary methods**

UKB contains linked Hospital Episode Statistics (HES) inpatient data for participants. The diagnosis table contains all diagnoses (ICD-10 code) associated with each admission episode. Individuals with a previous diagnosis of delirium were excluded to reduce potential bias through confounding and reverse causation. It also ensured all events occurred after baseline variables were measured at UKB enrolment and contributed to a more homogenous case-control cohort.

The operation table contains details of all OPCS4 procedure codes for a patient during a given admission. Inpatient surgery was defined according to patient class (excluding day case). Major surgery was defined by using a combination of two grading systems from the literature (Bupa schedule of procedures[21] and Abbott[20]). Eligible procedures were either Bupa ‘major’ or ‘complex’ category or, if not classified in Bupa, in the ‘restrictive’ category as defined by Abbott. Only procedures performed after enrolment in UK Biobank were eligible. All surgical specialties were included.

In cases of multiple procedures on the same date, the procedure coded as level 1 was treated as the index procedure. If there were multiple or zero level 1 procedures, the procedure with the highest grade was used (complex > major > restrictive). If there were multiple procedures with the highest grade, minimum array index was used as a tiebreak. Count variables were added to account for a) multiple surgical procedures within the same hospital admission and b) cumulative procedures since UKB enrolment.

A limitation of HES coding is that procedures have a specific date of operation, whereas diagnoses are linked to an entire inpatient episode (without a specific date of diagnosis). To limit the risk of a diagnosis preceding an operative procedure where both occurred in the same inpatient episode, the following decision tree was used:

Complication diagnosis in later episode than operation date?

Yes: include

No

Operation and diagnosis in same episode. Time from episode start to operation <14 days?

Yes

Operation date in first half of episode?

Yes: include

No: exclude

No: exclude
