## Supplementary material for "The genetic architecture of postoperative delirium after major surgery and its relationship with non-postoperative neurocognitive conditions": S3 Appendix

**Table A** Additional participant characteristics for case-control GWAS of postoperative delirium. Comorbidities assessed at time of surgery. Operative categories are listed as Abbott/Bupa classification.

| **Characteristic** | **Controls N = 139,148** | **Cases N = 1,016** |
| --- | --- | --- |
| Body mass index (kg/m2) | 27.5 [24.8-30.9] | 27.8 [25.0-31.2] |
| Missing | 716 (0.5%) | 8 (0.8%) |
| Townsend deprivation index | -2.1 [-3.6-0.6] | -1.8 [-3.4-1.2] |
| Missing | 158 (0.1%) | 1 (<0.1%) |
| Alcohol intake |  |  |
| Daily or almost daily | 27,891 (20%) | 287 (28%) |
| Three or four times a week | 29,888 (22%) | 185 (18%) |
| Once or twice a week | 35,295 (25%) | 232 (23%) |
| One to three times a month | 15,593 (11%) | 84 (8.3%) |
| Special occasions only | 17,787 (13%) | 118 (12%) |
| Never | 12,388 (8.9%) | 105 (10%) |
| Missing | 306 (0.2%) | 5 (0.5%) |
| Smoking status |  |  |
| Current | 15,070 (11%) | 158 (16%) |
| Previous | 53,340 (39%) | 448 (44%) |
| Never | 69,956 (51%) | 403 (40%) |
| Missing | 782 (0.6%) | 7 (0.7%) |
| Physical activity (days/week) | 4.00 [2.00-6.00] | 4.00 [2.00-7.00] |
| Missing | 8,779 (6.3%) | 88 (8.7%) |
| Myocardial infarction | 6,393 (5.2%) | 141 (14%) |
| Missing | 15,817 (11%) | 25 (2.5%) |
| Congestive cardiac failure | 3,189 (2.6%) | 137 (14%) |
| Missing | 15,817 (11%) | 25 (2.5%) |
| Cerebrovascular disease | 4,605 (3.7%) | 117 (12%) |
| Missing | 15,817 (11%) | 25 (2.5%) |
| Dementia | 375 (0.3%) | 56 (5.7%) |
| Missing | 15,817 (11%) | 25 (2.5%) |
| Chronic pulmonary disease | 15,504 (13%) | 261 (26%) |
| Missing | 15,817 (11%) | 25 (2.5%) |
| Diabetes | 9,527 (7.7%) | 170 (17%) |
| Missing | 15,817 (11%) | 25 (2.5%) |
| Diabetes with complications | 1,276 (1.0%) | 40 (4.0%) |
| Missing | 15,817 (11%) | 25 (2.5%) |
| Renal disease | 3,638 (2.9%) | 127 (13%) |
| Missing | 15,817 (11%) | 25 (2.5%) |
| Cancer | 16,303 (13%) | 223 (23%) |
| Missing | 15,817 (11%) | 25 (2.5%) |
| Operative category |  |  |
| Inclusive/complex | 418 (0.3%) | 4 (0.4%) |
| Inclusive/major | 138 (<0.1%) | 0 (0%) |
| Intermediate/complex | 6,726 (4.8%) | 28 (2.8%) |
| Intermediate/major | 18,734 (13%) | 40 (3.9%) |
| NOC/complex | 80 (<0.1%) | 0 (0%) |
| Restrictive/complex | 6,114 (4.4%) | 77 (7.6%) |
| Restrictive/major | 32,635 (23%) | 133 (13%) |
| Restrictive/NOC | 74,303 (53%) | 734 (72%) |

**Table B** Mapped genes from primary case-control GWAS of postoperative delirium. minGwasP represents the minimum p-value associated with the gene; IndSigSNPs lists the independent significant SNPs tagged to that gene.

| **Chr** | **Symbol** | **Type** | **minGwasP** | **IndSigSNPs** |
| --- | --- | --- | --- | --- |
| 19 | CTB-171A8.1 | antisense | 1.41254e-29 | rs12691088; rs429358; rs157582; rs11556505; rs75627662 |
| 19 | CTB-129P6.4 | antisense | 1.41254e-29 | rs11556505; rs157582; rs429358; rs10119; rs75627662; rs157592 |
| 19 | TOMM40 | protein_coding | 1.41254e-29 | rs11556505; rs157582; rs429358; rs10119; rs75627662 |
| 19 | APOE | protein_coding | 1.41254e-29 | rs10119; rs429358; rs11556505; rs75627662; rs12691088; rs157592; rs157582 |
| 19 | APOC1 | protein_coding | 1.41254e-29 | rs429358; rs11556505; rs75627662; rs12691088; rs157592; rs157582 |
| 19 | NKPD1 | protein_coding | 1.41254e-29 | rs429358; rs11556505; rs75627662 |
| 19 | APOC1P1 | pseudogene | 1.62817e-24 | rs157592; rs429358; rs12691088; rs75627662:rs429358; rs11556505; rs75627662 |
| 19 | AC084219.4 | antisense | 2.50438e-22 | rs11556505:rs429358; rs157582; rs11556505:rs157582:rs429358; rs157582:rs10119:rs75627662:rs429358; rs11556505; rs75627662:rs429358; rs75627662 |
| 19 | ZNF225 | protein_coding | 2.50438e-22 | rs12691088; rs11556505:rs429358; rs157582; rs11556505:rs157582:rs429358; rs157582:rs10119:rs75627662:rs429358; rs11556505; rs75627662:rs429358; rs75627662 |
| 19 | BCAM | protein_coding | 2.50438e-22 | rs11556505; rs429358; rs157582 |
| 19 | PVRL2 | protein_coding | 2.50438e-22 | rs12691088; rs11556505; rs157582; rs429358; rs10119; rs75627662; rs157592 |
| 19 | APOC4-APOC2 | protein_coding | 2.50438e-22 | rs10119:rs75627662:rs429358; rs11556505; rs75627662:rs429358; rs75627662:rs11556505:rs429358; rs157582; rs11556505:rs157582:rs429358 |
| 19 | APOC4 | protein_coding | 2.50438e-22 | rs10119:rs75627662:rs429358; rs11556505; rs75627662:rs429358; rs75627662:rs11556505:rs429358; rs157582; rs11556505:rs157582:rs429358 |
| 19 | APOC2 | protein_coding | 2.50438e-22 | rs10119:rs75627662:rs429358; rs11556505; rs75627662:rs429358; rs75627662:rs11556505:rs429358; rs157582; rs11556505:rs157582:rs429358 |
| 19 | CLPTM1 | protein_coding | 2.50438e-22 | rs429358; rs157582; rs11556505:rs157582:rs11556505:rs429358; rs157582:rs75627662:rs429358; rs11556505; rs75627662:rs429358; rs75627662:rs10119 |
| 19 | CLASRP | protein_coding | 2.50438e-22 | rs429358; rs157582; rs11556505:rs157582:rs11556505:rs429358; rs157582:rs75627662:rs429358; rs11556505; rs75627662:rs429358; rs75627662 |
| 19 | PVR | protein_coding | 1.39669e-20 | rs10119:rs75627662:rs429358; rs11556505; rs75627662:rs429358; rs75627662 |
| 19 | TRAPPC6A | protein_coding | 1.39669e-20 | rs75627662:rs429358; rs11556505; rs75627662 |
| 19 | BLOC1S3 | protein_coding | 1.39669e-20 | rs75627662:rs429358; rs11556505; rs75627662 |
| 19 | AC005779.2 | protein_coding | 1.39669e-20 | rs75627662:rs429358; rs11556505; rs75627662 |
| 11 | MS4A2 | protein_coding | 1.92752e-15 | rs429358; rs75627662 |

**Table C** Independent significant SNPs in non-cardiothoracic surgery case-control GWAS of postoperative delirium (sensitivity analysis). rsID: Reference SNP cluster ID.

| **rsID** | **Chr:position** | **p** | **Replication of primary GWAS?** |
| --- | --- | --- | --- |
| rs283815 | 19:45390333 | 4.24 x 10^-11^ | Proxy of rs157582, r^2^ 0.9759 |
| rs34404554 | 19:45395909 | 9.07 x 10^-12^ | Proxy of rs11556505, r^2^ 1.0 |
| rs10119 | 19:45406673 | 6.53 x 10^-9^ | Yes |
| rs429358 | 19:45411941 | 3.67 x 10^-20^ | Yes |
| rs157592 | 19:45424514 | 7.27 x 10^-16^ | Yes |

**Table D** Top 10 SNPs from case-control GWAS of postoperative delirium in cardiothoracic cohort (sensitivity analysis). rsID: Reference SNP cluster ID.

| **rsID** | **Chr:position** | **p** | **Independent significant SNP in primary GWAS?** |
| --- | --- | --- | --- |
| 19:45413234_GGT_G | 19:45413234 | 3.27 x 10^-7^ | No |
| rs429358 | 19:45411941 | 3.79 x 10^-7^ | Yes |
| rs12721051 | 19:45422160 | 6.11 x 10^-7^ | Proxy of rs157592, r^2^ 0.91 |
| rs56131196 | 19:45422846 | 6.52 x 10^-7^ | Proxy of rs157592, r^2^ 0.99 |
| rs283811 | 19:45388500 | 1.20 x 10^-6^ | Proxy of rs157582, r^2^ 0.99 |
| rs4420638 | 19:45422946 | 1.20 x 10^-6^ | Proxy of rs157592, r^2^ 0.93 |
| rs59007384 | 19:45396665 | 1.29 x 10^-6^ | Proxy of rs157582, r^2^ 0.85 |
| rs157582 | 19:45396219 | 1.56 x 10^-6^ | Yes |
| rs283815 | 19:45390333 | 1.85 x 10^-6^ | Proxy of rs157582, r^2^ 0.99 |
| rs157581 | 19:45395714 | 1.97 x 10^-6^ | Proxy of rs157582, r^2^ 0.99 |

**Table E** Independent significant single-nucleotide polymorphisms passing genome-wide significance in case-control GWAS of postoperative delirium after adjustment for age, sex, chip, first 10 PCs and Charlson Comorbidity Index (sensitivity analysis). rsID: Reference SNP cluster ID.

| **rsID** | **Chr:position** | **Nearest gene** | **Function** | **Effect allele** | **OR (95% CI)** | **p** |
| --- | --- | --- | --- | --- | --- | --- |
| rs429358 | 19:45411941 | *APOE* | exonic | T | 0.54 (0.49,0.6) | 5.5 x 10^-29^ |
| rs157592 | 19:45424514 | *APOC1* | intergenic | A | 0.58 (0.53,0.65) | 7.28 x 10^-23^ |
| rs157582 | 19:45396219 | *TOMM40* | intronic | C | 0.65 (0.59,0.72) | 7.55 x 10^-17^ |
| rs11556505 | 19:45396144 | *TOMM40* | exonic | C | 0.62 (0.56,0.7) | 1.02 x 10^-15^ |
| rs10119 | 19:45406673 | *TOMM40* | UTR3 | G | 0.7 (0.64,0.77) | 3.58 x 10^-13^ |
| rs75627662 | 19:45413576 | *APOE* | downstream | C | 0.72 (0.65,0.79) | 5.33 x 10^-10^ |
| rs12691088 | 19:45418486 | *APOC1* | intronic | G | 0.45 (0.35,0.57) | 1.28 x 10^-8^ |

**Table F** Polygenic risk scores from PGS Catalog passing matching and included in polygenic risk score analyses. *APOE-independent scores.

| **PGS Catalog Score ID** | **PGS Name** | **Number of Variants** | **Publication (doi)** |
| --- | --- | --- | --- |
| PGS000025* | GRS | 19 | 10.3233/JAD-150749 |
| PGS000026* | PHS | 31 | 10.1371/journal.pmed.1002258 |
| PGS000053 | ALZ21_NIA-LOAD | 21 | 10.1212/WNL.0000000000003734 |
| PGS000334 | GRSfull_22 | 22 | 10.1038/s41467-020-18534-1 |
| PGS000779 | PGS7_AD | 7 | 10.1002/dad2.12074 |
| PGS000811* | AD-PRS_39 | 39 | 10.1002/dad2.12142 |
| PGS000812* | AD-PRS_57 | 57 | 10.1002/dad2.12142 |
| PGS000823* | GRS23_AD | 23 | 10.1016/s1474-4422(18)30053-x |
| PGS000876* | PRS31_AD | 31 | 10.1002/acn3.716 |
| PGS000898* | PRS39_AD | 40 | 10.1038/s41467-021-22491-8 |
| PGS000945 | GBE_HC710 | 26 | 10.1371/journal.pgen.1010105 |
| PGS001349 | GBE_HC807 | 6 | 10.1371/journal.pgen.1010105 |
| PGS001775* | PRS39_AD | 39 | 10.1002/dad2.12229 |
| PGS001828 | portability-PLR_290.11 | 38 | 10.1016/j.ajhg.2021.11.008 |
| PGS002249 | AD_PRS_0.5 | 249273 | 10.1001/jama.2019.9879 |
| PGS002280* | GRS83_AD | 83 | 10.1038/s41588-022-01024-z |
| PGS002289 | GRS23_AD | 23 | 10.1001/jamanetworkopen.2022.5491 |
| PGS002731* | oA-PRS | 17 | 10.1212/wnl.0000000000200544 |
| PGS002753 | Alzheimer_s_disease_prscs | 1092011 | 10.1016/j.ajhg.2022.10.009 |
| PGS003440 | GRS11_nonapoeAD | 11 | 10.1038/s41598-023-28057-6 |
| PGS003441 | GRS28_AD | 28 | 10.1038/s41598-023-28057-6 |
| PGS003574 | GRS_Dementia21 | 21 | 10.1371/journal.pone.0277378 |
| PGS003953 | AD_Bellenguez | 1937 | 10.1186/s13195-023-01298-3 |
| PGS003954 | AD_FINNGEN | 81 | 10.1186/s13195-023-01298-3 |
| PGS003955 | AD_Jun | 85 | 10.1186/s13195-023-01298-3 |
| PGS003956 | AD_Kunkle_AFR | 157 | 10.1186/s13195-023-01298-3 |
| PGS003957 | AD_Kunkle | 12002 | 10.1186/s13195-023-01298-3 |
| PGS003958 | AD_Unweighted_PRSsum | 14109 | 10.1186/s13195-023-01298-3 |
| PGS003992 | dbslmm.auto.GCST90012877.AD | 1136212 | 10.1101/2023.11.20.23298215 |
| PGS004008 | lassosum.auto.GCST90012877.AD | 5663 | 10.1101/2023.11.20.23298215 |
| PGS004034 | ldpred2.auto.GCST90012877.AD | 1046908 | 10.1101/2023.11.20.23298215 |
| PGS004062 | megaprs.auto.GCST90012877.AD | 691136 | 10.1101/2023.11.20.23298215 |
| PGS004092 | prscs.auto.GCST90012877.AD | 1109233 | 10.1101/2023.11.20.23298215 |
| PGS004116 | pt_clump.auto.GCST90012877.AD | 58 | 10.1101/2023.11.20.23298215 |
| PGS004146 | sbayesr.auto.GCST90012877.AD | 915771 | 10.1101/2023.11.20.23298215 |
| PGS004227 | ad_apoe_gw_pgs | 15 | 10.1186/s13195-023-01184-y |
| PGS004228 | ad_apoe_0.1_pgs | 8863 | 10.1186/s13195-023-01184-y |
| PGS004229* | ad_noapoe_0.1_pgs | 8858 | 10.1186/s13195-023-01184-y |
| PGS004588* | PRS39_Eur | 39 | 10.1001/jamanetworkopen.2022.47162 |
| PGS004589* | PRS80_trans | 80 | 10.1001/jamanetworkopen.2022.47162 |
| PGS004590* | PRS363_rand_eff | 363 | 10.1038/s41380-023-02089-w |
| PGS004600* | PRS_AD83 | 83 | 10.1186/s12883-022-02925-6 |

**Table G** Odds ratio for postoperative delirium by Alzheimer’s disease polygenic risk score (PRS) quintile. Results are shown for all 42 polygenic risk scores, adjusted for age and sex, and the subset which did not include the APOE region (15 APOE-independent scores), adjusted for age, sex, APOE ε4 genotype and Charlson Comorbidity Index (0-1 vs >=2).

| **Term** | **All scores (n=42)** |  | ***APOE*-independent scores (n=15)** |  |
| --- | --- | --- | --- | --- |
|  | **Odds ratio (95% CI)** | **P value** | **Odds ratio (95% CI)** | **P value** |
| PRS quintile 1 | 1.00 (reference) | - | 1.00 (reference) | - |
| PRS quintile 2 | 1.08 (0.86,1.36) | 0.52 | 1.24 (1.00,1.53) | 0.05 |
| PRS quintile 3 | 1.27 (1.02,1.59) | 0.036 | 1.31 (1.06,1.62) | 0.01 |
| PRS quintile 4 | 1.48 (1.2,1.84) | <0.001 | 1.23 (1.00,1.53) | 0.05 |
| PRS quintile 5 | 2.29 (1.87,2.8) | <0.001 | 1.48 (1.2,1.82) | <0.001 |
| Age | 1.2 (1.19,1.22) | <0.001 | 1.2 (1.19,1.22) | <0.001 |
| Sex - male | 1.68 (1.47,1.92) | <0.001 | 1.68 (1.47,1.92) | <0.001 |
| *APOE* ε4 x1 | - | - | 1.74 (1.52,1.99) | <0.001 |
| *APOE* ε4 x2 | - | - | 4.13 (3.2,5.35) | <0.001 |
| CCI - >=2 | 2.89 (2.54,3.28) | <0.001 | 2.88 (2.53,3.27) | <0.001 |

**Table H** Results from primary analysis (case-control GWAS of postoperative delirium) for variants identified in candidate gene study by Heinrich et al. rsID: Reference SNP cluster ID.

| **Chr:position** | **rsID** | **Nearest gene** | **Effect allele** | **OR (95% CI)** | **P** |
| --- | --- | --- | --- | --- | --- |
| 11:46406767 | rs2067482 | CHRM2 | G | 0.98 (0.90,1.07) | 0.594 |
| 7:136701308 | rs8191992 | CHRM2 | T | 0.98 (0.90,1.07) | 0.64 |
| 7:136701935 | rs6962027 | CHRM4 | T | 0.95 (0.84,1.08) | 0.426 |

**Figure A** Manhattan plot for case-control GWAS of postoperative delirium after previous dementia diagnoses excluded. The red line represents a genome-wide significant p value of 5 x 10^-8^.

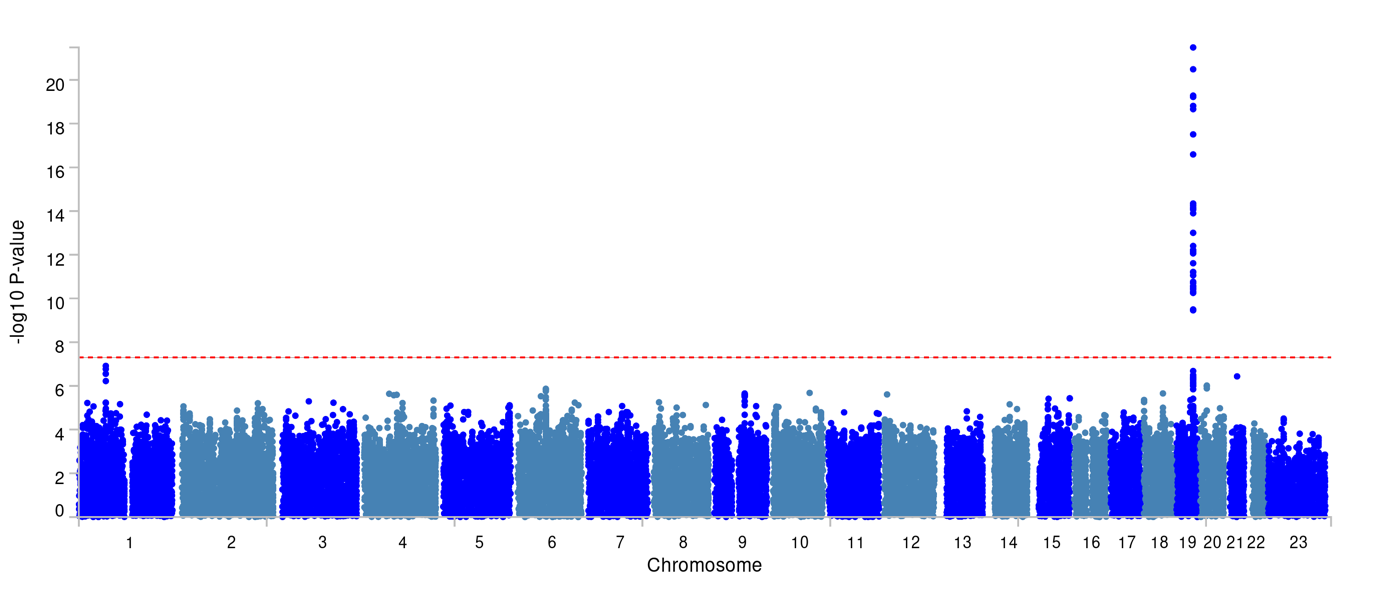

**Figure B** Manhattan plot for MAGMA testing for case-control GWAS of postoperative delirium after previous dementia diagnoses excluded

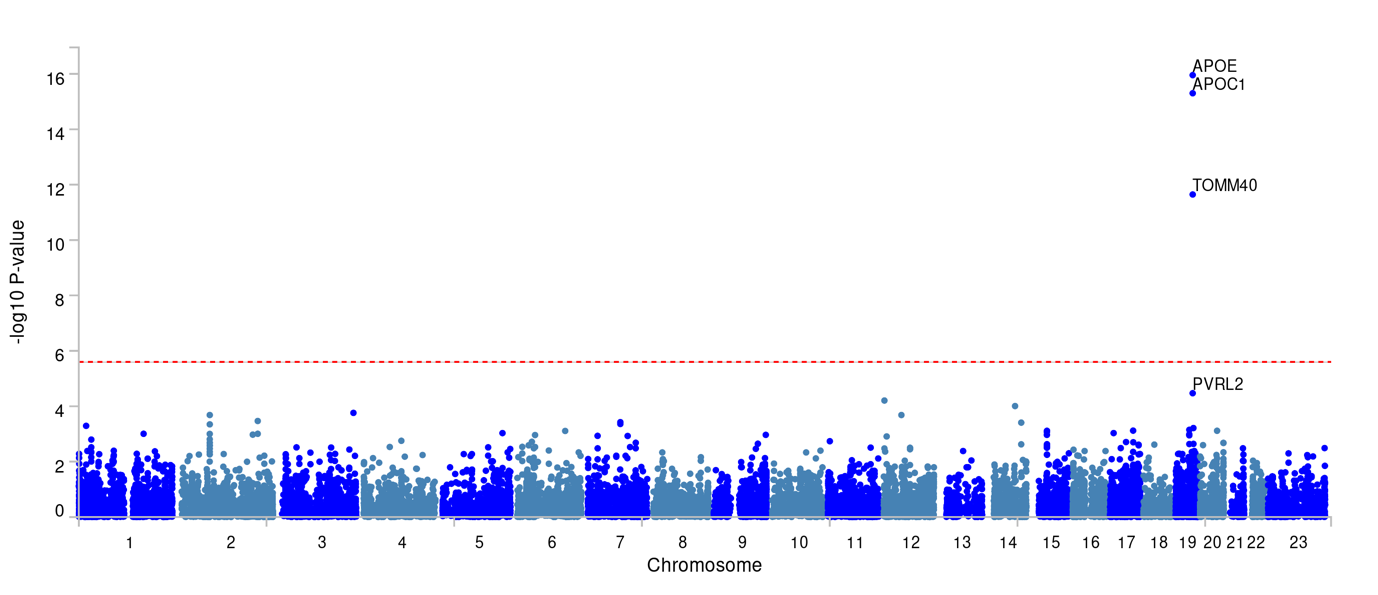

**Figure C** Manhattan plot for case-control GWAS of postoperative delirium after previous or subsequent dementia diagnoses excluded. The red line represents a genome-wide significant p value of 5 x 10-8.
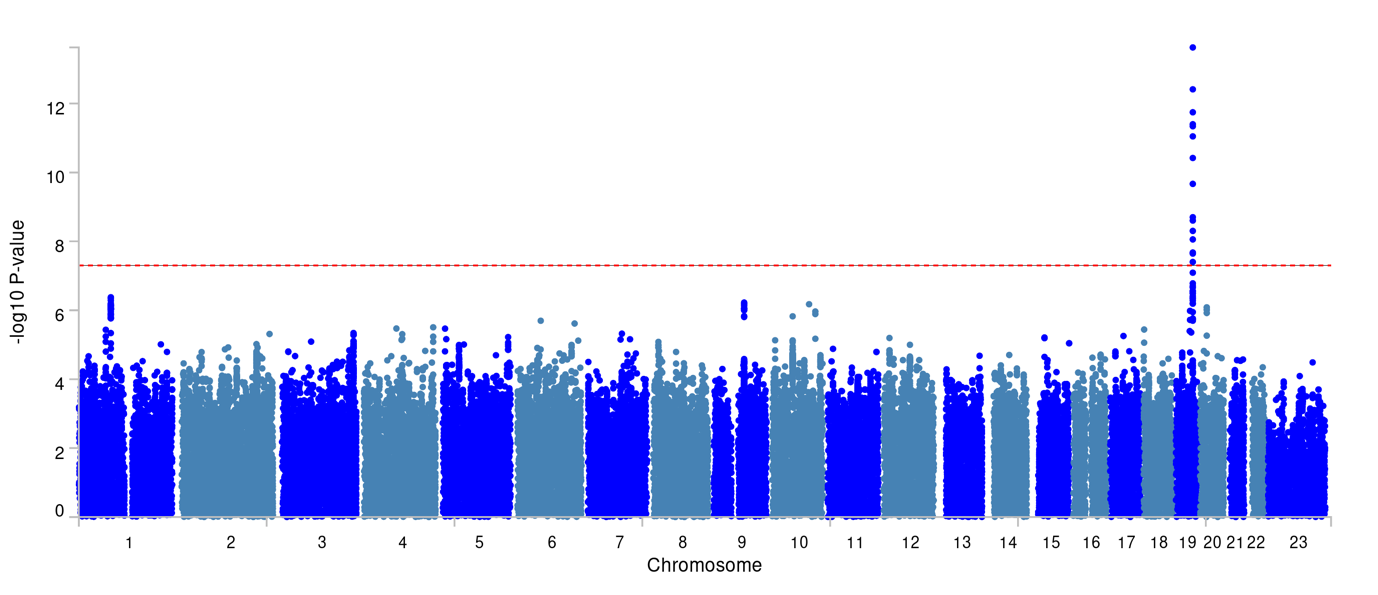

**Figure D** Manhattan plot for MAGMA testing for case-control GWAS of postoperative delirium after previous or subsequent dementia diagnoses excluded

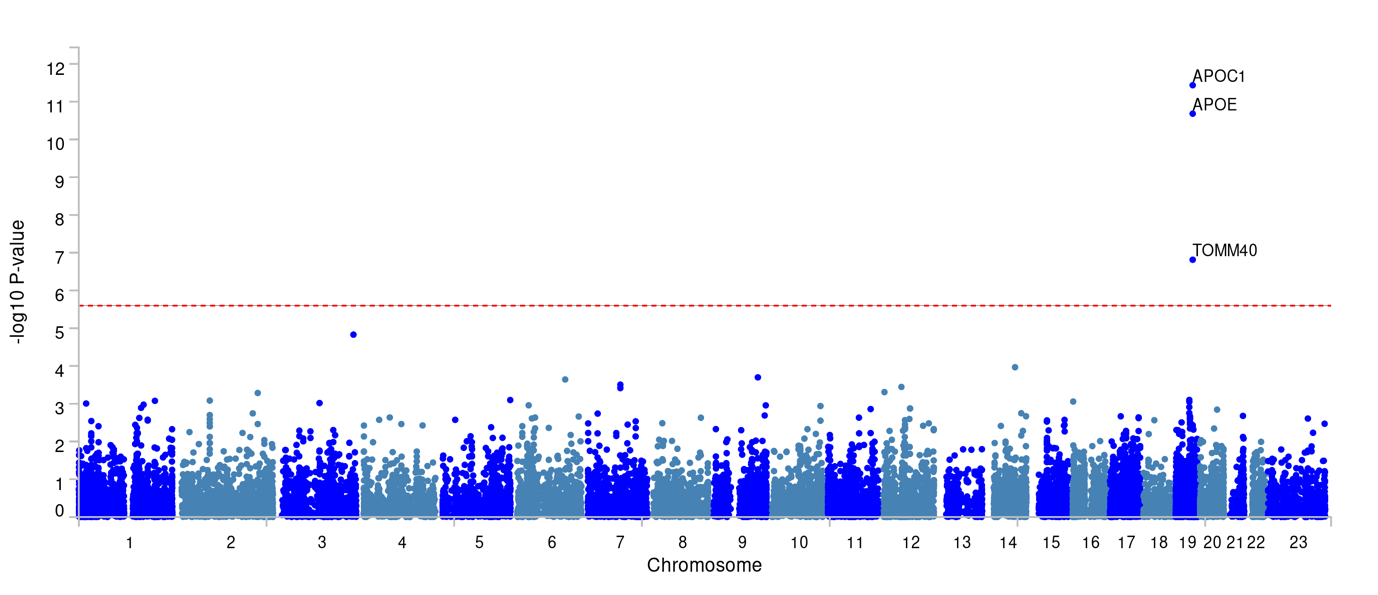

**Figure E** Manhattan plot for case-control GWAS of postoperative delirium conditional on APOE ε4 allele count (adjusted for age, sex, chip, first 10 PCs)

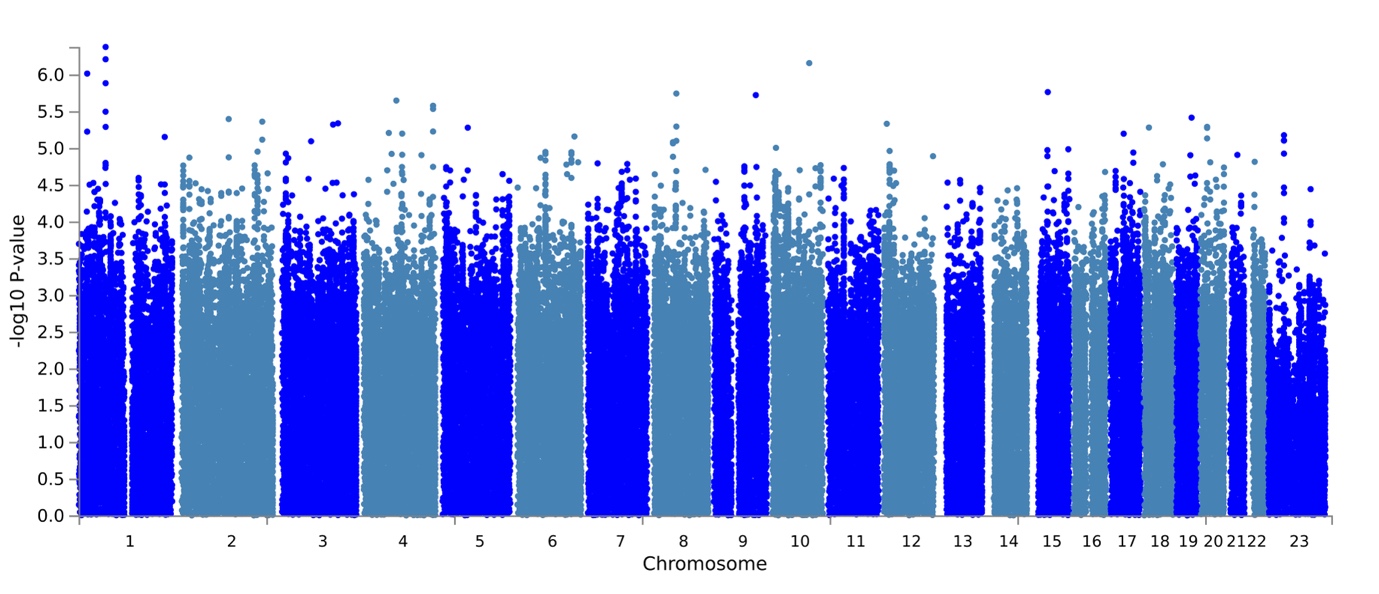

**Figure F** GWAS catalog reported genes for case-control GWAS of postoperative delirium (primary analysis)

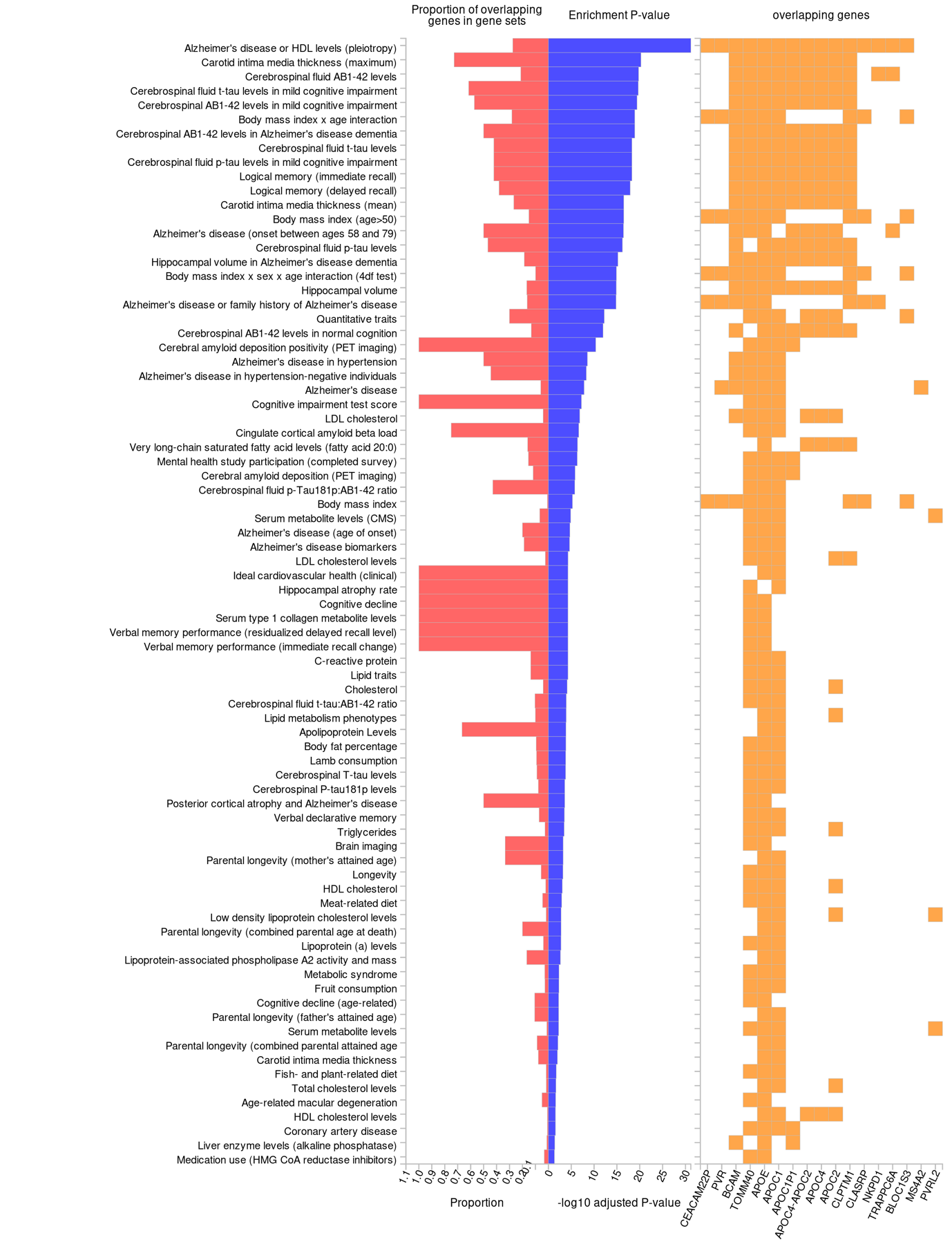
